# Rapid CRISPR-based surveillance of SARS-CoV-2 in asymptomatic college students captures the leading edge of a community-wide outbreak

**DOI:** 10.1101/2020.08.06.20169771

**Authors:** Jennifer N. Rauch, Eric Valois, Jose Carlos Ponce-Rojas, Zach Aralis, Ryan S. Lach, Francesca Zappa, Morgane Audouard, Sabrina C. Solley, Chinmay Vaidya, Michael Costello, Holly Smith, Ali Javanbakht, Betsy Malear, Laura Polito, Stewart Comer, Katherine Arn, Kenneth S. Kosik, Diego Acosta-Alvear, Maxwell Z. Wilson, Lynn Fitzgibbons, Carolina Arias

**Affiliations:** University of California Santa Barbara, Department of Molecular, Cellular, and Developmental Biology; Neuroscience Research Institute, University of California, Santa Barbara; Student Health Service, University of California Santa Barbara; Department of Pathology, Santa Barbara Cottage Hospital; Pacific Diagnostic Laboratories, Santa Barbara; Department of Medical Education and Division of Infectious Diseases, Santa Barbara Cottage Hospital; Center for BioEngineering, University of California, Santa Barbara; Center for Stem Cell Biology and Engineering, University of California, Santa Barbara

## Abstract

**Importance:** The re-opening of colleges and universities through the US during the COVID-19 pandemic is a significant public health challenge. The development of accessible and practical approaches for SARS-CoV-2 detection in the college population is paramount for deploying recurrent surveillance testing as an essential strategy for virus detection, containment, and mitigation.

**Objectives:** In this study, we set out to determine the prevalence of SARS-CoV-2 in asymptomatic subjects in a university community, using, for the first time, CREST, a CRISPR-Cas13-based test that we developed for accessible and large scale viral surveillance^1^.

**Study Design, Setting, and Participants:** We enrolled 1,808 asymptomatic persons to undergo SARS-CoV-2 testing. We compared viral prevalence in self-collected oropharyngeal swab samples obtained in two time periods: May 28th-June 11^th^ and June 23rd-July 2nd, 2020. We detected viral genomes in these samples using CREST, and we corroborated our results with a point-of-reference RT-qPCR test. Positive samples were confirmed by a clinical laboratory and reported to the local Public Health Department.

**Results:** All the 732 tests performed between late May to early June were negative. In contrast, tests performed on 1,076 samples collected between late June to early July revealed eight positive cases by CREST, confirmed by RT-qPCR and CLIA-diagnostic testing. The average age of the positive cases was 21.7 years; all individuals self-identified as students. These metrics showed that CREST was effective at capturing positive SARS-CoV-2 cases in our student population. Notably, the viral loads detected in these asymptomatic cases resemble those seen in clinical samples, highlighting the potential of covert viral transmission.

**Conclusions and Relevance:** Our study revealed a substantial shift in SARS-CoV-2 prevalence in a young and asymptomatic population, and uncovered the leading wave of a local outbreak that coincided with rising case counts in the surrounding county and the state of California. Moreover, and most notably, the almost perfect concordance between CRISPR- and PCR-based testing indicated that CREST using self-collected OP swabs is reliable and it allows expanding options for large-scale surveillance testing and detection of SARS-CoV-2 outbreaks, as is required to resume operations in higher education institutions in the US and abroad.

**Key Points:** *Question:* Are CRISPR-based methods a reliable and accessible option to capture SARS-CoV-2 outbreaks in a college population?

*Findings:* We tested 1,808 asymptomatic individuals from a university population for SARS-CoV-2, using, for the first time, a CRISPR-based assay for virus surveillance. We detected eight positive cases, corroborated by RT-qPCR and confirmed by a clinical laboratory. Our CRISPR-based method captured a change in viral prevalence coinciding with the relaxation of lockdown measures and the rise of COVID-19 cases in the community.

*Significance:* We demonstrate that CRISPR-based methods offer scalable and reliable SARS-CoV-2 testing for virus surveillance and allow capturing the leading edge of an outbreak.

## Background

The COVID-19 pandemic has claimed hundreds of thousands of lives and has disrupted the way of life of countless communities. To control this pandemic, communities worldwide closed businesses, prohibited large social gatherings, and adopted non-pharmacological intervention (NPI) measures^2–4^. Initial restrictions were successful in several countries where COVID-19 cases, hospitalizations, and deaths declined^3,4^. However, as communities relaxed social distancing and restrictions, COVID-19 cases returned, often following exponential growth. Several metrics, including percent positivity of testing, hospitalizations, and death rates, have been used to gain insights into epidemic trends in specific populations. Prevalence among asymptomatic persons is an important but more elusive metric, primarily because of test scarcity and prioritization of symptomatic patients or contacts with confirmed cases. Nevertheless, understanding both asymptomatic prevalence and the impact of NPI measures on infection rates has tremendous potential to inform vital public health decisions.

A fundamental aspect of pandemic control is careful planning for the re-opening of college campuses. While COVID-19 testing has focused on individuals with increased risk of infection and mortality, an increasing disease burden has emerged in those aged 19-30, many of whom attend college and university^5^. Every year since 2017, over 15 million students attend colleges in the US^6^. Many students reside in dormitories and off-campus housing, frequently in crowded conditions, sharing restrooms, kitchens, and common areas^7^. These living conditions are associated with high morbidities of diseases like meningococcal meningitis, influenza, mumps, and measles^8–11^. Respiratory pathogens like SARS-CoV-2 are easily transmitted among individuals living in college dormitories and during social contact by exposure to live virus in aerosol droplets^12–14^.

Further complicating SARS-CoV-2 transmission in university settings is the well-documented infectivity of asymptomatic persons, many of whom are likely to be pre-symptomatic with high viral loads^15–21^. Those without symptoms are likely to be responsible for as many as 44% of new infections^22^. Recent examples of colleges re-opening and promptly closing, or those that implemented drastic quarantine measures for their students following the detection of COVID-19 outbreaks, illustrate the challenges of safely bringing academic activities back to campus during a pandemic. The upsurge of cases within college populations also presents a risk beyond campus walls, as infections can spill over to neighboring communities^23^. The early identification of infected individuals through expanded and frequent surveillance testing is essential to curb disease spread. However, before undertaking such large-scale surveillance testing, the prevalence of asymptomatic infection must be ascertained to inform decisions regarding the utility of expanded testing in a university population^24^.

To understand viral prevalence in the university community, and to assess the potential of a CRISPR-based test to screen for SARS-CoV2 in asymptomatic persons, we enrolled healthy volunteers from the University of California, Santa Barbara (UCSB) in a virus surveillance study. We obtained self-collected oropharyngeal (OP) swab samples, which we processed for SARS-CoV-2 testing using two methods; CREST, our newly-developed CRISPR-Cas13-based assay^1^, and the CDC-recommended RT-qPCR assay^25^, which we used as a point-of-reference test. We compared the results obtained from two time periods. The first collection period occurred during May-June, 2020, approximately two months into a state-wide stay-at-home mandate. The second collection period occurred during late June-early July 2020, approximately three weeks after local restrictions for isolation were relaxed in the community. Our results revealed no COVID-19 cases in our study population in May-June of 2020. Using the same methods, we demonstrated a substantial shift in prevalence approximately one month later, which coincided with changes in community restrictions and public interactions. Notably, CREST performed as well as the CDC-recommended RT-qPCR assay. Our study substantiates, for the first time, the utility of self-collected OP swabs and CRISPR-based testing as valuable alternatives for large-scale surveillance sampling of SARS-CoV-2 in asymptomatic individuals.

## Methods

### Study population

UCSB’s population includes 26,134 students and 5,668 staff and faculty. 38% of the students live in university housing, and 34% in the nearby community of Isla Vista (23,096 residents, 1.866 mi^2^, 12,377 people/mi^2^). This study was open to all symptom-free individuals, 18 years of age or older, affiliated with UCSB (student, faculty, staff, direct relatives). Individuals who exhibited a fever (100.4°F), cough, or shortness of breath in the two weeks before or on the day of sample collection were excluded from the study. Only five subjects were excluded due to presenting symptoms at the time of collection and were referred to local healthcare resources.

### Sample collection

UCSB healthcare professionals collected informed consent and demographic data (age, address, telephone, gender, and UCSB affiliation) at the sampling locale. Samples were assigned a numeric code for deidentification purposes. Samples were acquired as self-collected OP swabs stored in PBS, with surveillance by a healthcare professional. Samples were inactivated at 56 °C for 30 minutes, and RNA was extracted using the QIAamp MinElute Virus Spin Kit (Qiagen 57704) or Viral RNA Mini Kit (Qiagen 52906) from 140-200 µL of sample, and eluted in 50 µL. Pre- and post-analytical protocols were reviewed and approved by the Santa Barbara Cottage Hospital IRB.

### SARS-CoV-2 detection by RT-qPCR (one-step TaqMan assay)

Viral RNA was reverse transcribed and amplified using the TaqPath one-step cDNA master mix kit (ThermoFisher 501148245) following the manufacturer’s recommendations. Reactions were prepared as previously described^1^. Briefly, a 15 μL master mix reaction was prepared using the established CDC protocol^26^, and 5 μL of RNA were added into the reaction with each of the target-specific TaqMan primers and probes. For no template controls, 5 μL of nuclease-free water were used. Positive control reactions used 10^6^ copies of *in vitro* transcribed RNA encoding the SARS-CoV-2 nucleocapsid sites N1 and N2. Reactions were run the BioRad CFX96 Touch qPCR instrument with the thermal profile: 25 °C/2 minutes; 50 °C/15 minutes; 45 cycles of 95 °C/5 seconds followed by 55 °C/30 seconds and plate read; hold at 4 °C. Data were analyzed using the BioRad CFX Maestro software using a single threshold for Cq determination. We prepared standard curves of *in vitro* transcribed RNAs, ranging from 10^6^ to 10^0^ copies/μL, to determine detection limits. One-way ANOVA with a post-hoc Dunnett’s test was used to determine the Cq value significance from no template control using Prism v8 software (Graphpad). The limit of detection for N1 and N2 is 10^2^ copies/µL (Cq of 32.59 and 34.405, respectively) and for RNaseP 10^3^ copies/µL (Cq = 34.328). Samples were considered positive if the signal for both N1 and N2 was above the limit of detection.

### CREST (Cas13-based, Rugged, Equitable, Scalable Testing)

CREST reactions were carried out as described^1^. Briefly, 5 µL of RNA were reverse transcribed using RevertAid (200 U/μL, ThermoFisher Scientific) in the presence of murine RNase inhibitor (NEB). Water was used as negative control. Positive control reactions used 10^6^ copies of *in vitro* transcribed RNA. The reaction mixtures were heated to 42 °C/30 minutes, then placed on ice. 2 µL of the resulting cDNAs was used as templates for PCR amplification with *Taq* DNA polymerase (NEB), using the thermal profile: 98 °C/2 minutes; 20 cycles of 98 °C/15 seconds, 60 °C/15 seconds, and 72° C/15 seconds; final extension at 72°C/5 min. Cas13a was used for site-specific detection using fluorescent probes. The reaction was performed in Cas13a cleavage buffer (40 mM Tris pH 7.5, 1 mM DTT) supplemented with 1 mM rNTPs (ThermoFisher Scientific), 2 U/μL RNase Inhibitor (NEB), 0.125 μM cleavage reporter (IDT), 1.5 U/μL T7 RNA Polymerase (Lucigen), 6.3 ng/μL LwaCas13a, 20 nM Cas13 crRNA and 9 mM MgCl_2_. Reactions were composed of 4 μL Cas13a cleavage solution and 1 μL of sample (RT-PCR product) in a well of a 384 well-plate, with samples run in duplicate or quadruplicate wells. Fluorescence was acquired every 5 minutes for 30 minutes at 37 °C in a Quantstudio5 qPCR instrument (Applied Biosystems). The initial reading taken at time = 0 was subtracted from time = 30 to get a ΔRFU for each well. To determine a threshold for negative and positive results, ΔRFU from negative control wells were multiplied by 5 and used as a cutoff. Plates were valid if negative control reactions did not increase 3x during the experiment. Samples were considered positive if the signal for both N1 and N2 was 5X above the background.

### Primer, gRNA and cleavage reporter sequences

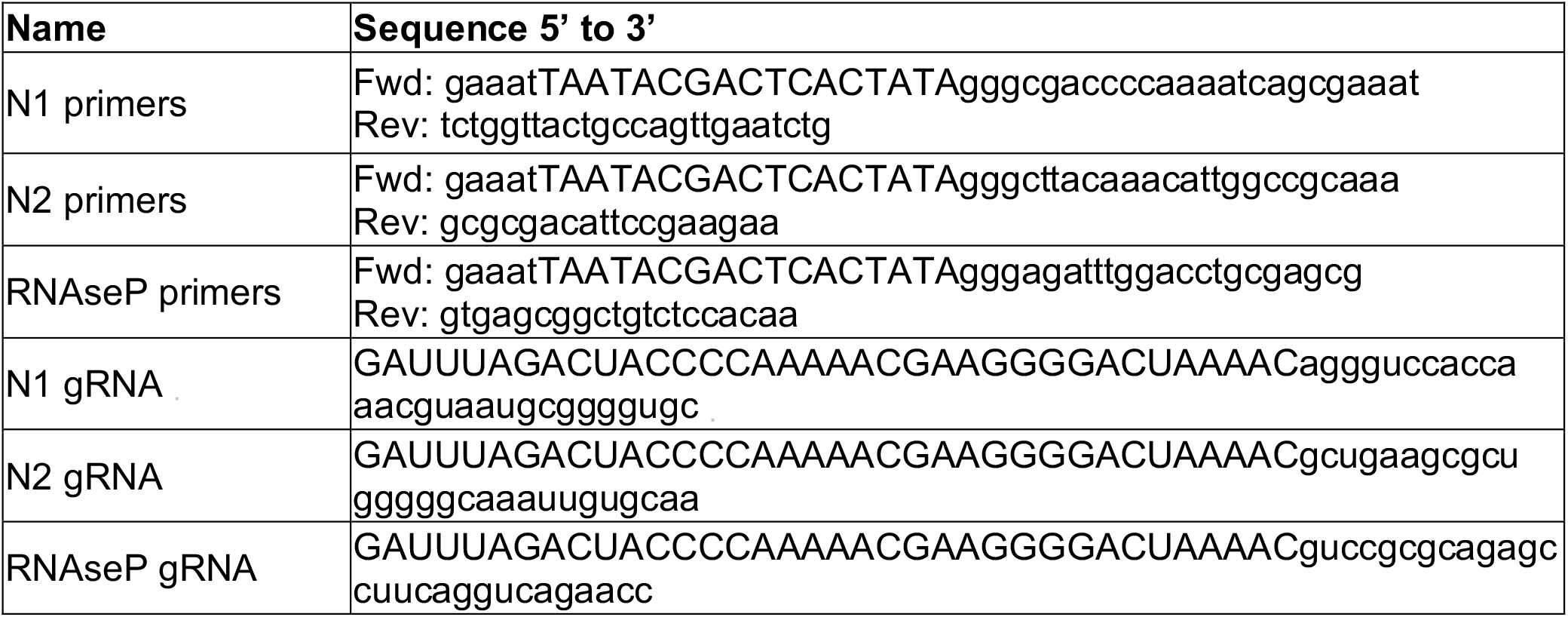
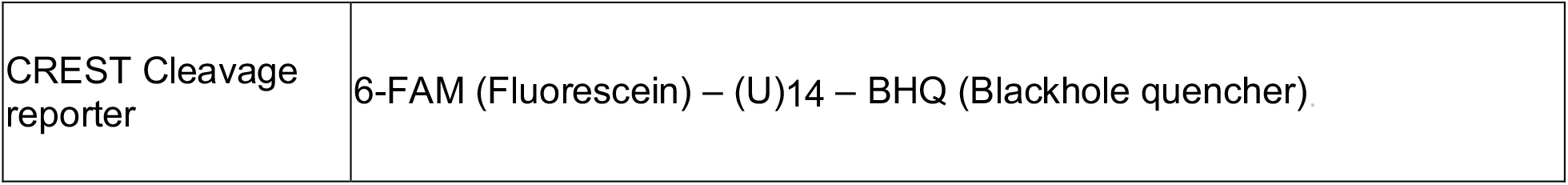

### Analyses

Correlations between N1 and N2 and between CREST and TaqMan assays were calculated using the Pearson correlation coefficient, assuming data are from a bivariate normal distribution, using the R function cor.test(). Percent positive rates (shown in figure 4) were fit using a logistic growth model where 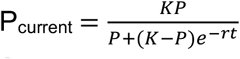, with K = 100%, P = 0.03, and r fit by minimizing the error found to be r = 0.101.

**Figure 1.**
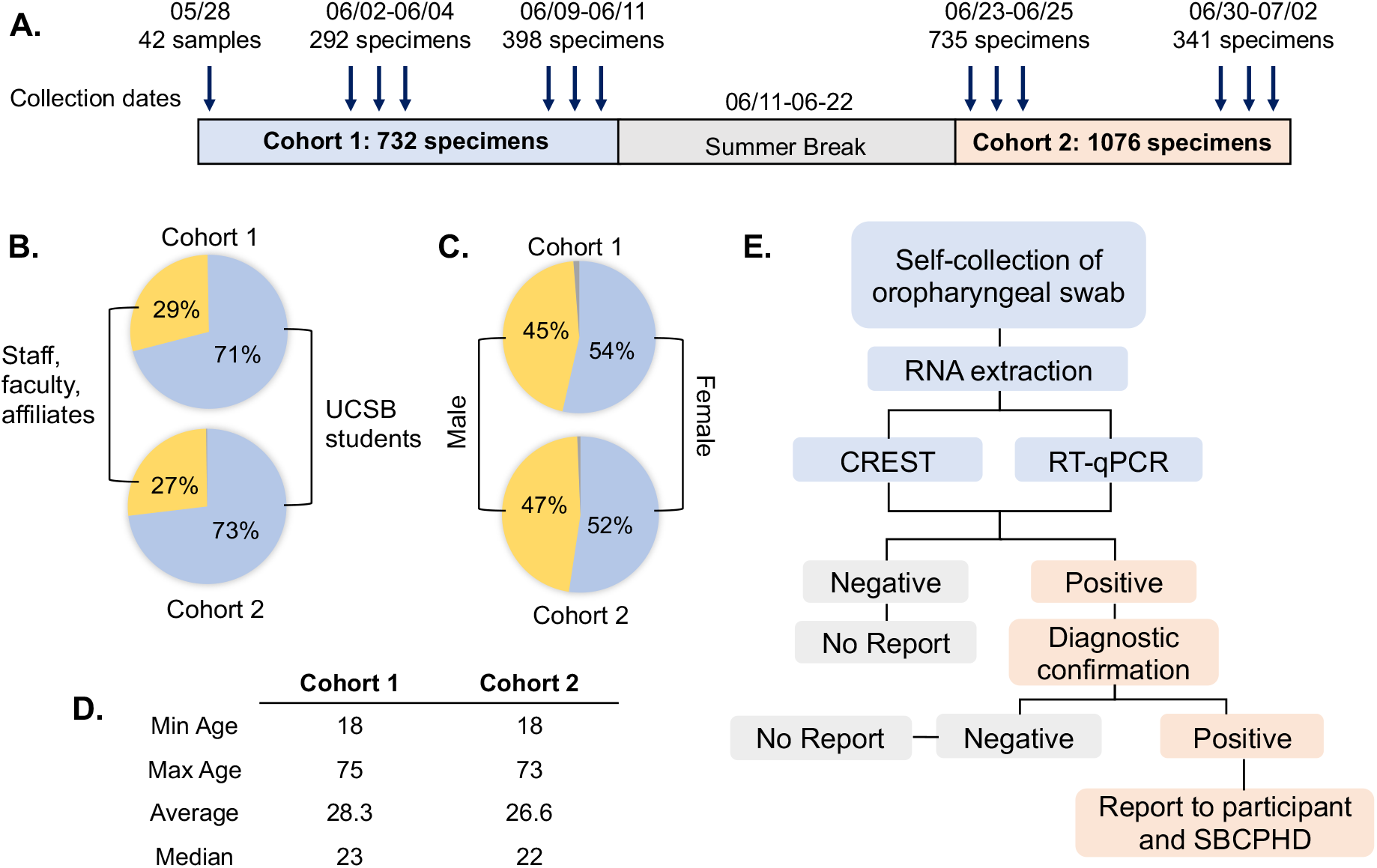
Description of the study and population demographics. (A) For this study, we recruited 1,808 asymptomatic subjects in two cohorts during May, June, and July of 2020. The arrows indicate the days of sample collection. We did not collect samples during the UCSB summer break from June 11 to June 22. (B, C, D) Demographics of the study population, including UCSB affiliation (B), gender (C), and age of study participants (D). (E) Flow chart of specimen handling. We processed self-collected OP swabs for SARS-CoV-2 testing using CREST or RT-qPCR. We handled samples downstream from testing according to the result. We did not report negative results to the participants. We submitted positive results for confirmation with diagnostic testing to a CLIA laboratory. Following confirmation, Santa Barbara Cottage Hospital Clinicians reported the positive results to the participants and the Santa Barbara County Public Health Department (SBCPHD).

**Figure 2.**
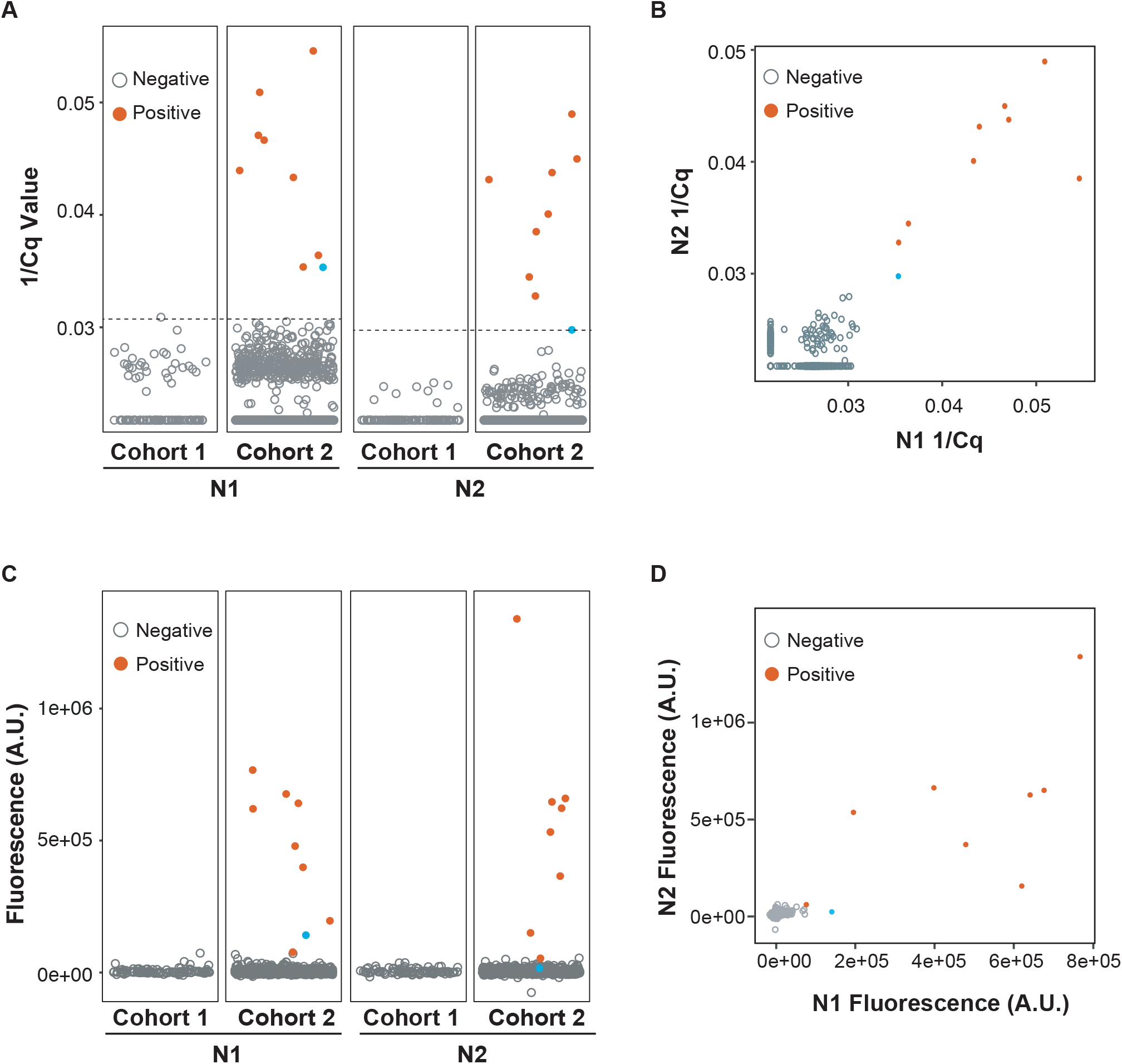
Detection of positive samples by RT-qPCR and CREST. (A) Distribution of the 1/Cq values from RT-qPCR by cohort. (B) correlation of N1 and N2 signal detected by RT-qPCR. (C) Distribution of the fluorescence values from CREST by cohort. (D) correlation of N1 and N2 signal by CREST. Grey open dots indicate negative samples; solid red dots indicate positive samples. The blue dot indicates one sample detected by RT-qPCR, but not confirmed by CREST or in a CLIA laboratory test. (Note the low level of N2 by CREST for this sample). The dashed line in panel (A) indicates the detection limit for RT-qPCR (N1 1/Cq 0.0306, N2 1/Cq 0.029).

**Figure 3.**
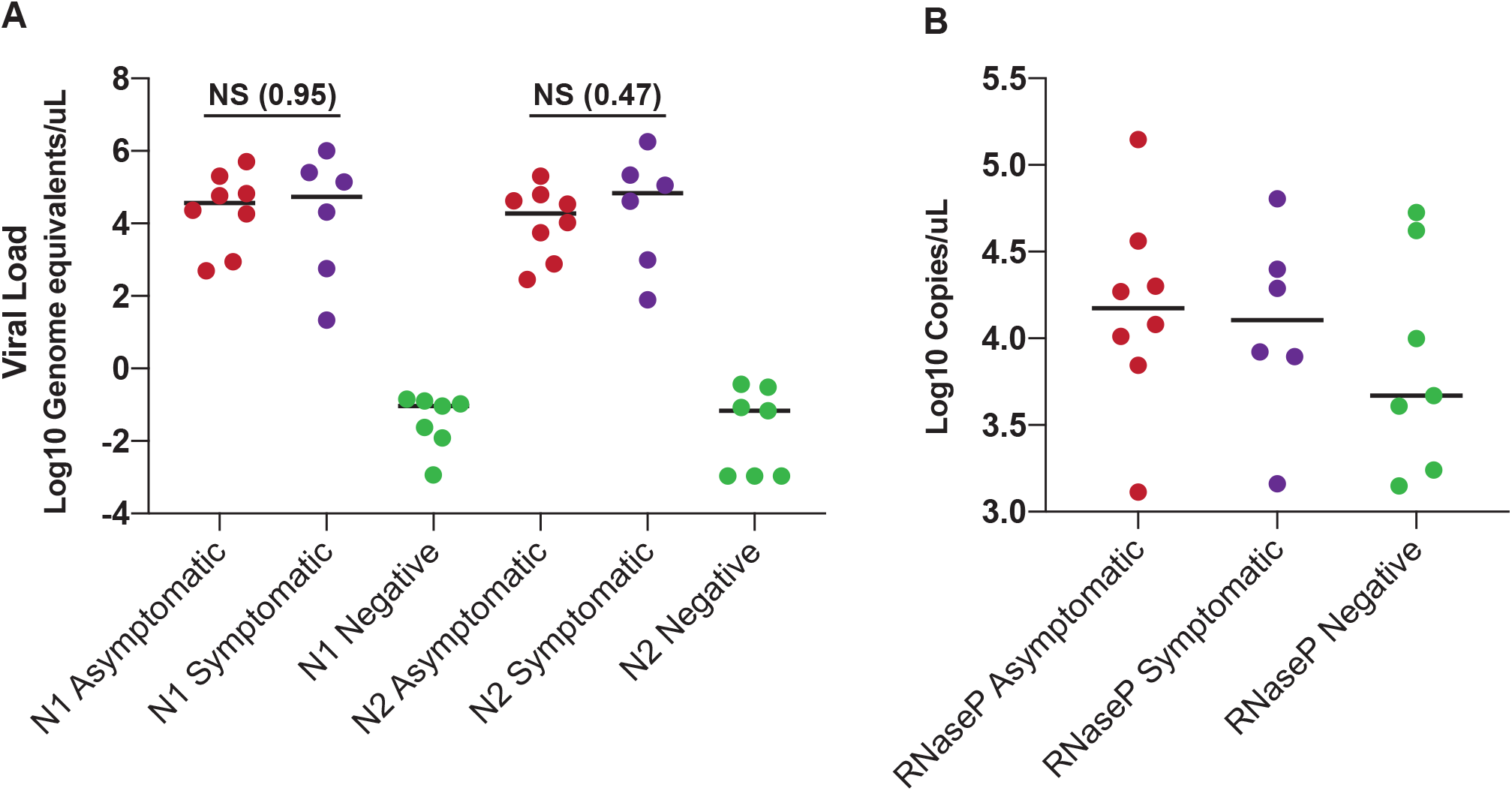
Viral loads in asymptomatic and confirmed positive individuals. (A) Viral loads, expressed as genome equivalents/µL, were calculated using the RT-qPCR data for N1 and N2 detection. (B) RNase P copies were calculated using the RT-qPCR data for this host gene target. In our analyses, we included the eight positive samples we detected in cohort 2, that were confirmed by diagnostic testing (red). As controls we included residual clinical samples from known positive (purple, N = 6) or negative patients (green, N = 7) provided to us by our collaborators at the SBCPHD. (A) Median, solid line. NS: p = 0.95 for N1, p = 0.497 for N2 Mann-Whitney test. (B) Median, solid line. NS: p = 0.63 for RNase P Kruskal-Wallis test.

**Figure 4.**
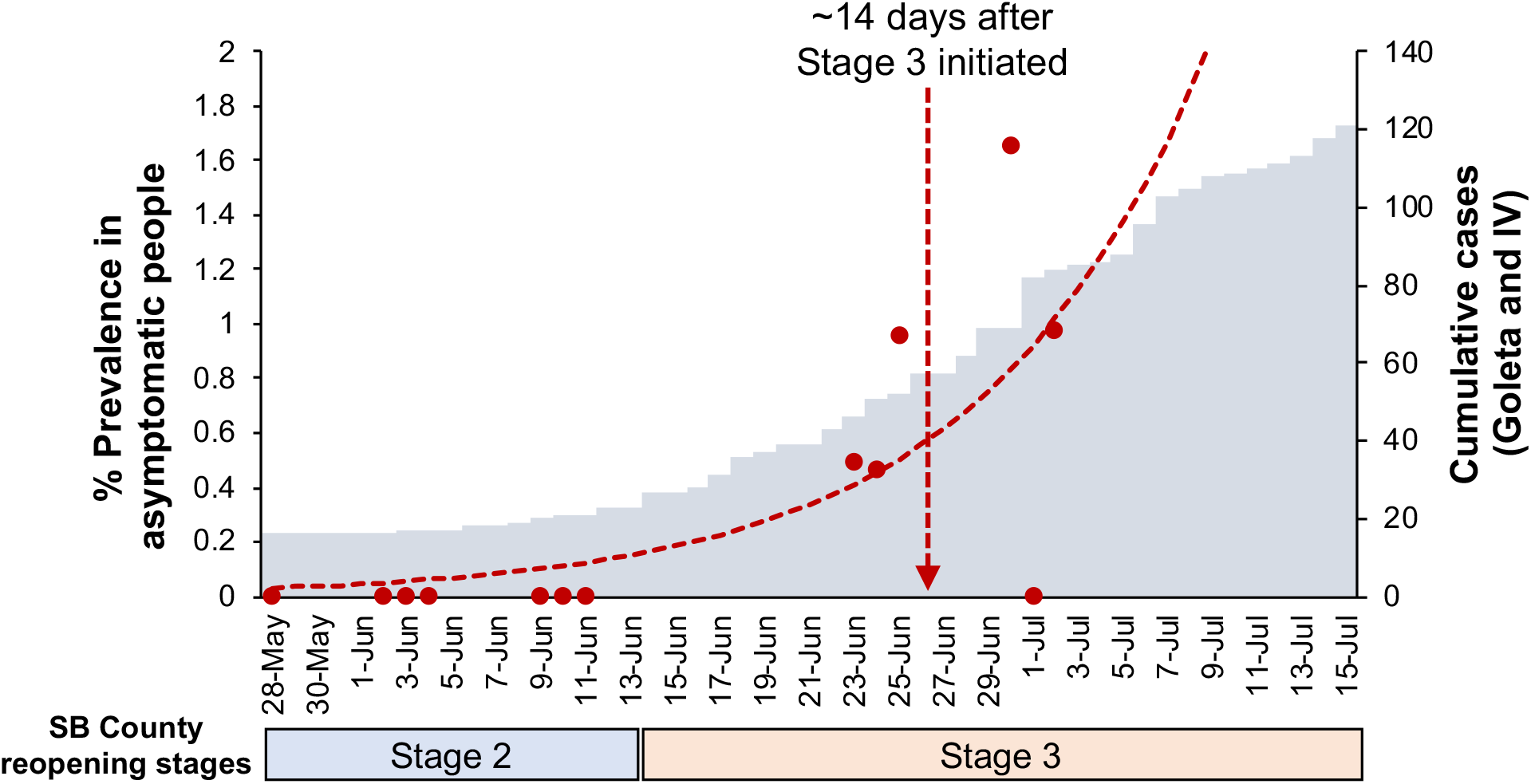
Dynamics of SARS-CoV-2 prevalence in the study population. We calculated the daily viral prevalence for the samples in this study as a percent of positive cases (red dots). We calculated the trendline, indicated by the red dotted line, by finding the *r* in a logistic growth model that minimized the error while fixing the percent prevalence on May 28 to 0.03%. The grey bars represent the cumulative daily number of diagnosed COVID-19 cases in the Goleta and Isla Vista communities, based on official data from the SBCPHD. The timing of stage two and three of the California re-opening plan in Santa Barbara county are below the graph.

## Results

To obtain insights on the prevalence of SARS-CoV-2 in our local community, we enrolled 1,808 healthy volunteers in a surveillance study. All participants were asymptomatic for COVID-19 at the time of sample collection. Among the participants, 1,805 reported affiliation with UCSB, and 1,306 (72.2%) were undergraduate and graduate students (Sup. Fig. 1A). This population reflects the composition of the UCSB community (Sup. Fig. 1A).

We acquired self-collected OP swabs from the participants over two time periods, from May 28 to June 11 (Cohort 1), and from June 23 to July 2 (Cohort 2) (Fig. 1A). Over 70% of the subjects in both cohorts self-identified as UCSB students (71% cohort 1; 73% cohort 2) (Fig. 1B), 45-47% were male, 52-54% were female (Fig. 1C), and 67% of all participants reported the UCSB neighboring communities of Goleta and Isla Vista as their place of residence (Sup. Fig. 1B). Our study population’s average age was 28.3 and 26.6 years old for cohort 1 and 2, respectively, with a minimum age of 18 years old and a maximum of 73-75 years old (Fig 1D and Sup. Fig. 1C).

We used two assays to detect SARS-CoV-2 genomes in the OP swab samples: CREST, a CRISPR-Cas13-based method we recently developed^1^, and the RT-qPCR test recommended by the CDC^25^, which we used as a point-of-reference (Sup. Fig. 2). Both methods detect two sites in the nucleocapsid gene, N1 and N2, and one site in the host RNase P transcript, which ensured consistency in our analyses. Samples were processed in-house with a turnaround time from 12-30 hours from the moment of collection. All samples collected in cohort 1 (N = 732) were negative by both tests (Fig. 2A, C). In contrast, we detected eight positive samples by CREST and nine by RT-qPCR in cohort 2 (N = 1,076) (Fig. 2A, C). We found a good correlation in detecting the nucleocapsid gene using the N1 and N2 primers (RT-qPCR, Pearson correlation coefficient r = 0.566, Fig 2B) and probes (CREST, Pearson correlation coefficient r = 0.872, Fig. 2D). The positive subjects’ average age was 21.7 years old, and all self-identified as UCSB students (Sup. Table 1). The eight samples detected by CREST were independently confirmed by a CLIA-certified laboratory test, and the results were reported to the participants and the Santa Barbara County Public Health Department (SBCPH) by Santa Barbara Cottage Hospital (SBCH) clinicians (Fig. 1E). One sample was positive solely by RT-qPCR at the detection threshold, reflecting a low viral copy number (Sup. Table 1, Sup. Table 3). With this single possible exception, RT-qPCR and CREST results were concordant (Sup. Fig. 3).

The participants with positive tests were offered the opportunity to follow up with clinicians at the UCSB student health service (SHS). Six out of eight individuals provided an update of symptoms to the UCSB SHS. Two subjects reported no symptoms, two subjects reported mild symptoms (nasal congestion, sore throat), and two subjects reported classic COVID-19 symptoms (fatigue, anosmia). None of the participants reported fever as a symptom (Sup. Table 2).

To estimate the viral load in the asymptomatic or pre-symptomatic subjects confirmed as positive, we calculated the genome equivalents per µL based on the Cq values for N1 and N2 from the RT-qPCR assay, using linear regression on a standard curve ranging from 10^0^ to 10^6^ gene copies/uL. Our study subjects’ viral load ranged from 286 to 510,000 copies/µL (Sup. Table 3). These viral load levels were not significantly different from those detected in a control set of de-identified residual nasopharyngeal (NP) swab samples obtained from symptomatic patients in the local community and provided to us by our collaborators at the SBCPHD (Sup. Table 3, Fig. 3). Notably, the quality of the self-collected specimens using OP swabs was not significantly different from those collected using NP swabs (positive and negative controls), as measured by the detection of RNase P transcripts (p = 0.63, Kruskal-Wallis test) (Fig. 3).

Next, we calculated the prevalence of SARS-CoV-2 in our study population using the confirmed cases. The viral prevalence of cohort 1 was 0%, while that of cohort 2 was 0.74%, with a daily incidence ranging from 0 to 1.65% (Fig. 4, Sup. Table 4). The change in prevalence between cohorts was statistically significant (p = 0.013, Fisher’s exact test). The prevalence dynamics in our study population reflect the increase in COVID-19 cases diagnosed in the UCSB neighboring communities of Goleta and Isla Vista, where 67% of our participants reside (Fig .4). The increase in the number of infections detected in our study—and those in Santa Barbara County—coincided with the implementation of stage three of the California re-opening plan in Santa Barbara County (Fig. 4, Sup. Fig. 5).

## Discussion

As colleges and universities through the US struggle to recover from the academic, social, and economic impacts of months of remote learning, a pressing trial remains: how to re-open campuses safely? A primary challenge for university communities is the potential for covert infections promoted by social and academic gatherings, which are unavoidable in the context of a vibrant university campus. Recent evidence indicates that asymptomatic and pre-symptomatic individuals can unknowingly transmit the virus and fuel covert outbreaks^20,27,28^. The early detection of asymptomatic infections, particularly those with high SARS-CoV-2 loads like those detected in our analyses, which may underlie super-spreader events, is vital for mitigating viral transmission and containing outbreaks. This information is also essential to guide university directives to make decisions regarding campus opening across the country and ensure superior education continuity. Epidemiological models support this notion and suggest that universal and frequent SARS-CoV-2 testing is necessary for efficient disease containment^24^. However, the economic impact of providing reliable and regular testing for thousands of students, faculty, and staff may prohibit larger campuses from closely monitoring their communities.

With the considerations above in mind, we evaluated for the first time the performance of our recently developed CRISPR-based strategy for large-scale viral surveillance in asymptomatic subjects. This method, known as CREST, uses PCR amplification and Cas13 for the detection of viral genomes with a simple binary outcome. CREST is as efficient at detecting SARS-CoV-2 infections in asymptomatic subjects as the CDC recommended RT-qPCR, which is considered the “gold standard” testing method. This CRISPR-based method also has the added benefit of enabling an easy to interpret and dependable binary readout, fluorescence vs. no fluorescence. CREST showed perfect concordance with positive cases diagnosed in a CLIA certified laboratory (Pacific Diagnostics Laboratory), further corroborating its robustness. Because we designed CREST to be a low-cost and accessible method, it offers a much-sought alternative for communities where resources are limited, and where access to testing is difficult. Besides, CREST is scalable, enabling high throughput testing, and it uses laboratory-generated or off-the-shelf commercially available reagents, thus eliminating the restriction of limiting supply chains. For these reasons, we surmise CREST can offer a solution for places where access to professional laboratories is restrictive and instances in which a high volume of repetitive sampling is necessary, including the university setting.

In addition to validating CREST as an alternative for surveillance testing, we also evaluated the use of supervised self-collected OP swabs as the source sample for asymptomatic surveillance. Self-collection methods minimize the risk of exposure for health care personnel and maximize sample acquisition efficiency. Our data support that self-sampling by OP swab is dependable, and thus provides an alternative method for unsupervised or remotely supervised sample acquisition outside of a healthcare setting. OP and other self-sampling methods could enable sample processing by mail, which can substantially enhance testing coverage^29,30^.

One of our most significant observations is the difference in SARS-CoV-2 prevalence between the two cohorts we analyzed. While we did not detect any infections in the 732 people tested in May/June, approximately one month later, we demonstrated a shift in prevalence, with eight confirmed cases among 1,076 asymptomatic people surveyed. This significant change in the transmission dynamics coincided with the release of community restrictions and increased public and social interactions during the implementation of stage three of the California re-opening plan in Santa Barbara County. The increase in prevalence was exclusive to young and asymptomatic individuals (average age 21.7 years old, range 19-30 years old) who self-identified as UCSB students, and who may not otherwise have accessed COVID-19 testing. Individuals in this age group are likely to be socially active, highlighting how easily covert infections could result in flare-ups. Our surveillance program detected the initial wave from a local outbreak and coincided with rising case counts in the Goleta and Isla Vista localities, the Santa Barbara County, and the State of California.

Overall, our study provides strong evidence supporting the use of CRISPR-based assays and self-collected OP swabs and as feasible, rapid, and dependable tools for the surveillance of SARS-CoV-2 in asymptomatic individuals. The concordance between RT-qPCR testing and our strategy of using OP swabs and CREST substantiates the feasibility of using simpler, equally robust approaches for high-volume, recurrent testing, which is a desirable strategy to facilitate the re-opening of colleges and universities. Monitoring the population to detect COVID-19 cases before they lead to outbreaks could constitute the paramount containment and mitigation approach within large campus communities and others facing similar challenges.

## Data Availability

All data referred to in the manuscript, will be made available upon request

## Acknowledgments

We thank the UC Santa Barbara Office of Research for their generous support. We thank Dr. Mary Ferris and the UCSB Student Health Service personnel who helped establish the pre-analytical protocols and collect the samples. We thank Laura Isaac, Erin Ross, and Catelynn Kenner at SBCH for their guidance on the IRB protocols. We thank all the participants in our study. Finally, we thank all essential workers for keeping society running. Without them, this work would have never happened.

## Figures and Figure Legends

**Supplementary Figure 1.**
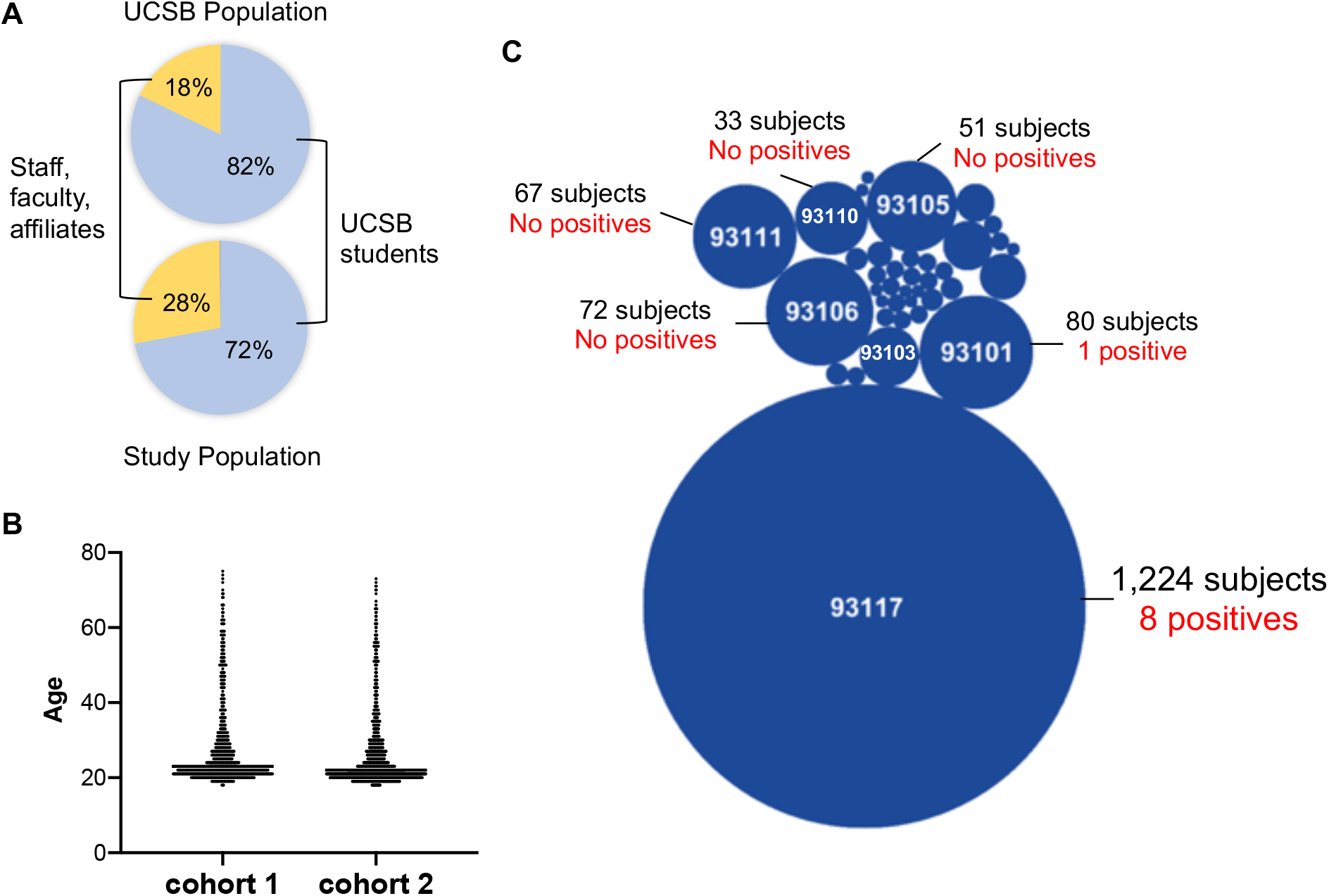
Study population demographics. (A) Affiliations of the UCSB population. (B) Distribution of zip codes reported by the participants. The size of the bubble reflects the size of the population from that zip code. (C) Distribution of the age of participants by cohort.

**Supplemental Figure 2.**
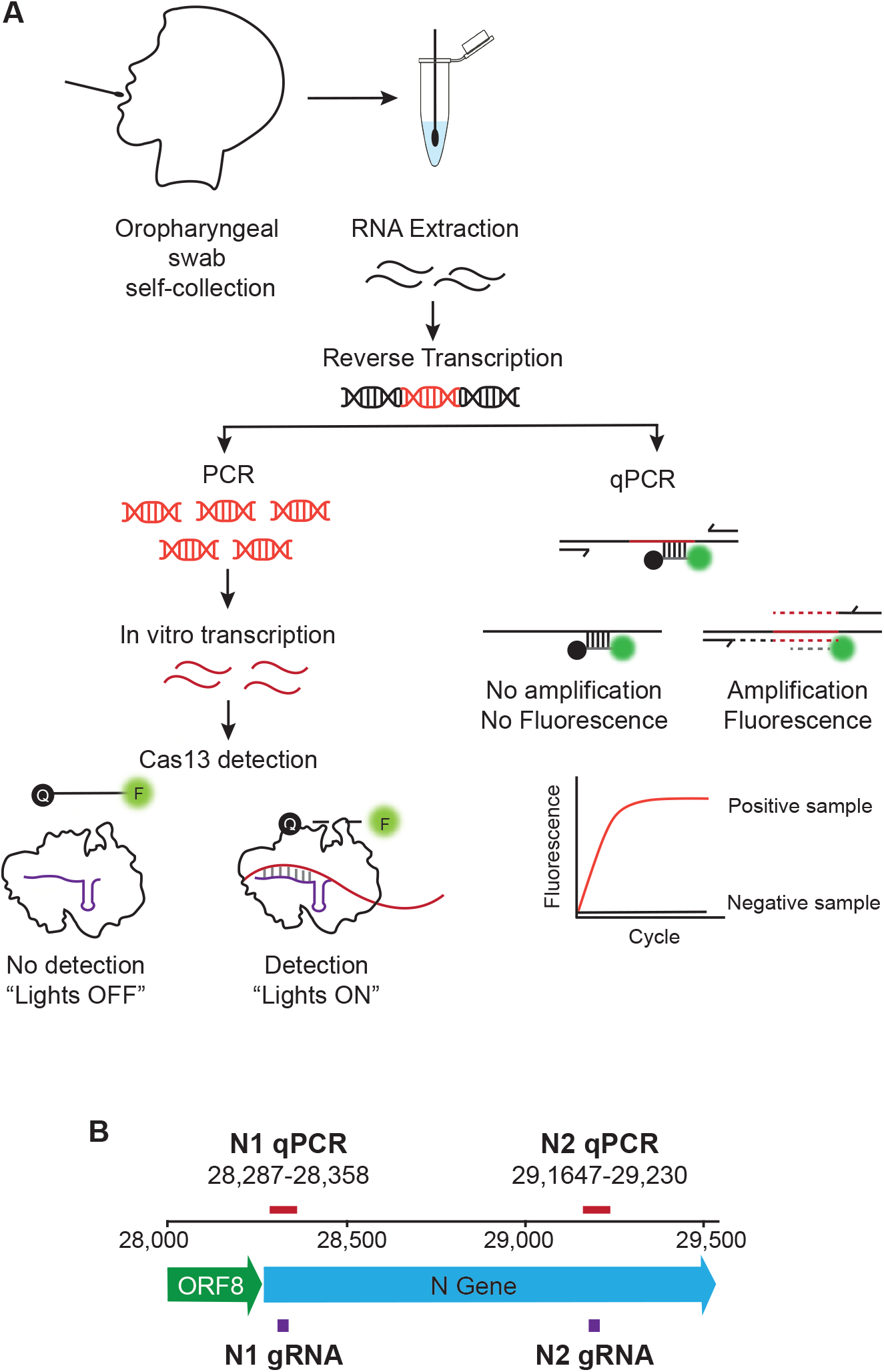
Overview of CREST and RT-qPCR protocols. (A) The participants collected OP swabs, supervised by a healthcare provider. We extracted the RNA from the samples and handled it according to the method to use for testing. For CREST, the RNA was reverse transcribed, and the resulting DNA was amplified by polymerase chain reaction (PCR) using primers for the N1, N2, and RNAse P target regions (see panel B). The amplified region of interest was transcribed *in vitro* and used as the template for detection by Cas13. The activation of Cas13 following target recognition by the guide RNA (gRNA) was measured using a fluorescent poly-U cleavage reporter. For qPCR, the RNA was reverse transcribed and detected by real-time amplification. (B) Genomic map of the SARS-CoV-2 genome regions detected in this study.

**Supplementary Figure 3.**
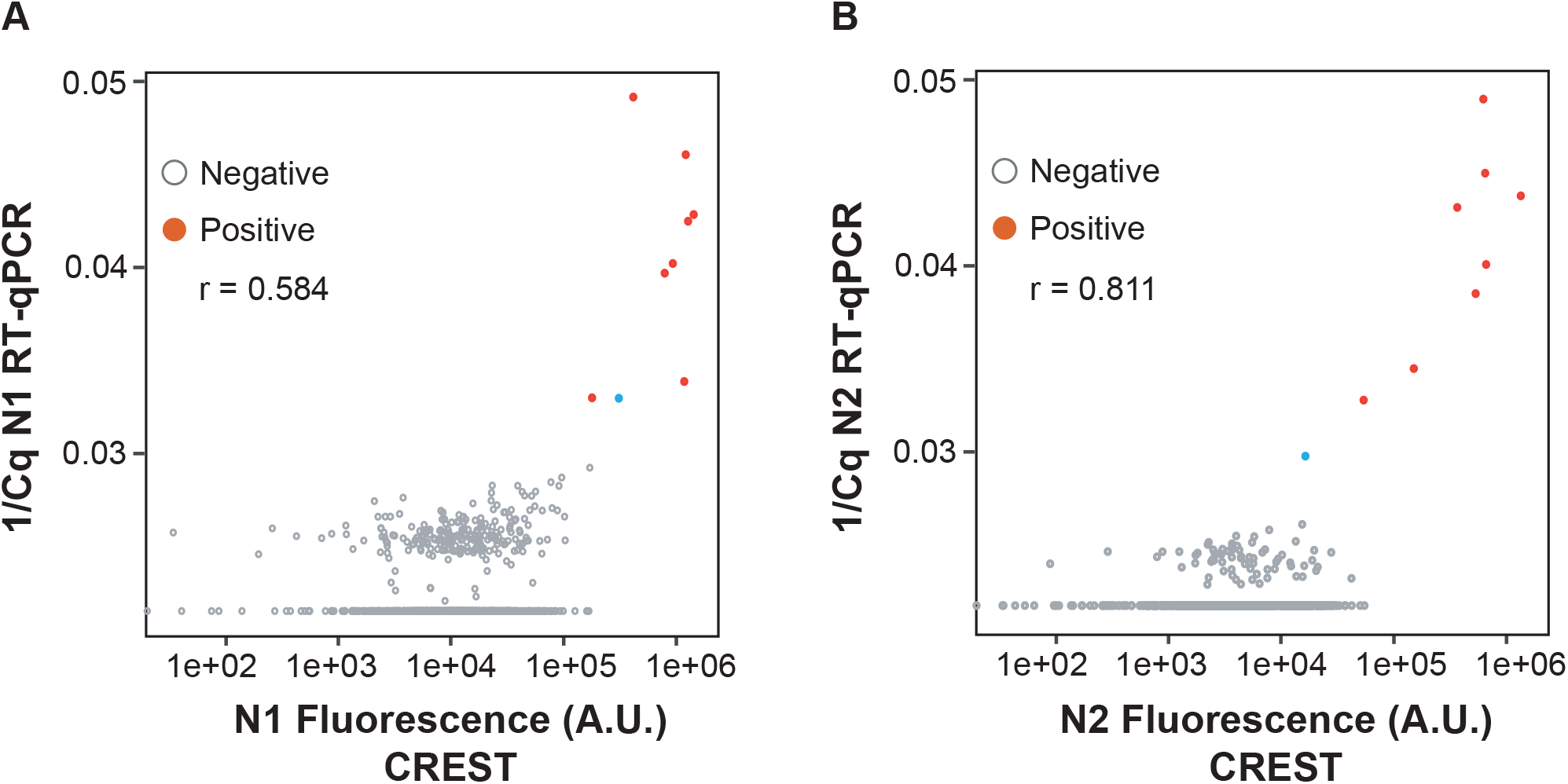
Correlation between RT-qPCR and CREST detection of positive and negative samples. Correlation of the signal detected for N1 (A) or N2 (B) in RT-qPCR and CREST. Grey open dots indicate negative samples; solid red dots indicate positive samples. The blue dot indicates one sample detected by RT-qPCR, but not confirmed by CREST or in a CLIA laboratory test. Pearson correlation coefficient N1 r = 0.584, N2 r = 0.811.

**Supplementary Figure 4.**
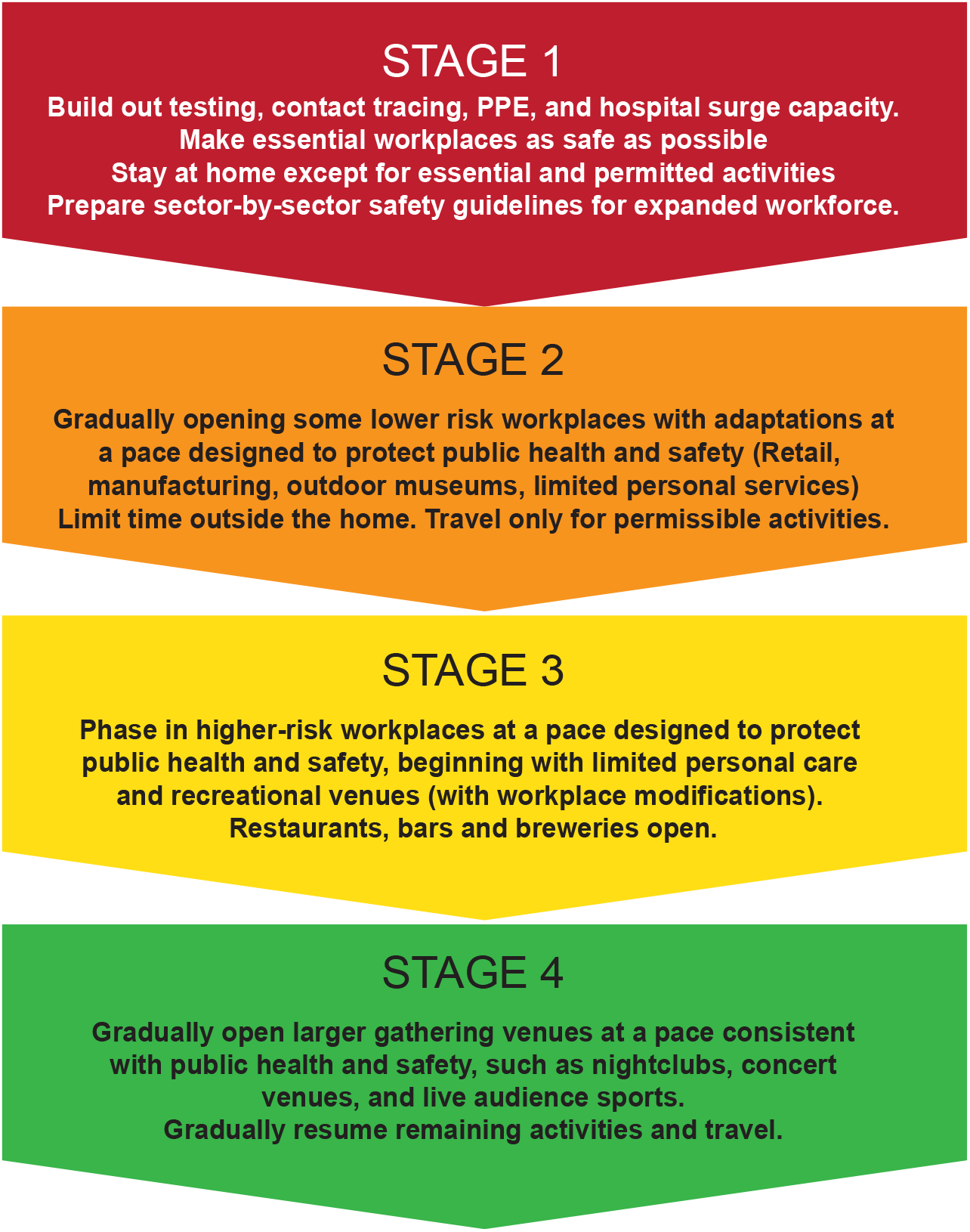
Description of the California re-opening plan stages. Our study took place during the transition between stage 2 (Low-risk workplaces) and stage 3 (higher-risk workplaces) https://covid19.ca.gov/roadmap/.

**Supplementary Table 1.** Age of participants, CREST fluorescence signal, and RT-qPCR Cq values for each of the positive samples detected in the study. The negative control is the average signal for all no template sample. The positive control is the average signal for all samples where we used in vitro transcribed RNA as a template. The N2 signal for sample 4 was below the level of detection in CREST, and close to the limit of detection by RT-qPCR. This sample was not confirmed as positive by a diagnostic test performed in a CLIA certified laboratory.

| Sample | Date of collection | Age | CREST (Fluorescence) |  |  | RT-qPCR (Cq) |  |  |
| --- | --- | --- | --- | --- | --- | --- | --- | --- |
|  |  |  | N1 | N2 | RNAseP | N1 | N2 | RNAse P |
| 1 | 6/23/20 | 20 | 767040 | 1339679 | 678959 | 21 | 23 | 28 |
| 2 | 6/24/20 | 22 | 620196 | 150156 | 730279 | 27 | 29 | 28 |
| 3 | 6/25/20 | 20 | 676364 | 646462 | 629386 | 21 | 22 | 23 |
| *4 | 6/25/20 | 20 | 141479 | 16355 | 613756 | 28 | 34 | 26 |
| 5 | 6/25/20 | 21 | 641187 | 622513 | 458237 | 20 | 20 | 28 |
| 6 | 6/25/20 | 23 | 398685 | 659525 | 345808 | 23 | 25 | 31 |
| 7 | 6/30/20 | 21 | 196032 | 532165 | 470316 | 18 | 26 | 29 |
| 8 | 6/30/20 | 30 | 479062 | 365208 | 710805 | 23 | 23 | 27 |
| 9 | 7/2/20 | 19 | 77259 | 53825 | 450471 | 28 | 31 | 25 |
| Neg. Cont. | N/A | N/A | 7741 | 5980 | 5980 | 43 | 43 | 43 |
| Pos. Cont. | N/A | N/A | 243665 | 441170 | 371064 | 11 | 11 | 18 |

**Supplementary Table 2.** List of the symptoms that were reported by participants with positive SARS-CoV-2 tests in this study. Following CLIA-confirmation of their SARS-CoV-2 status, positive individuals SBCH clinicians contacted the participants and provided them with recommendations to follow, including the opportunity to contact the UCSB Student Health Service clinicians. Six out of eight positive individuals contacted the UCSB SHS clinicians and reported symptoms. None of the participants reported fever as a symptom.

| Outcome | Number of cases | Symptoms reported |
| --- | --- | --- |
| Asymptomatic | 2 | None |
| Pre-symptomatic (mild) | 2 | Nasal congestion, sore throat |
| Pre-symptomatic (classic) | 2 | Fatigue, anosmia |

**Supplementary Table 3.** Viral load (genome equivalents/µL), calculated for the positive samples in this study, and the known positive and negative samples from the community of Santa Barbara County. Sample 4 was not confirmed by diagnostic testing in a CLIA certified laboratory.

| Viral load |  |  |  |
| --- | --- | --- | --- |
| Sample | Date of collection | N1 (gen. equ./μL) | N2 (gen. equ./μL) |
| 1 | 6/23/20 | 67000 | 42000 |
| 2 | 6/24/20 | 871 | 760 |
| 3 | 6/25/20 | 58500 | 62700 |
| *4 | 6/25/20 | 488 | 38 |
| 5 | 6/25/20 | 204000 | 204000 |
| 6 | 6/25/20 | 18600 | 10600 |
| 7 | 6/30/20 | 510000 | 5500 |
| 8 | 6/30/20 | 23000 | 34000 |
| 9 | 7/2/20 | 500 | 286 |
| Pos. Cont. 1 | N/A | 1006971 | 1781555 |
| Pos. Cont. 2 | N/A | 139311 | 112919 |
| Pos. Cont. 3 | N/A | 254744 | 214151 |
| Pos. Cont. 4 | N/A | 571 | 989 |
| Pos. Cont. 5 | N/A | 20866 | 41588 |
| Pos. Cont. 6 | N/A | 21 | 79 |
| Neg. Cont. 1 | N/A | 0.107 | 0.001 |
| Neg. Cont. 2 | N/A | 0.024 | 0.068 |
| Neg. Cont. 3 | N/A | 0.127 | 0.368 |
| Neg. Cont. 4 | N/A | 0.001 | 0.302 |
| Neg. Cont. 5 | N/A | 0.093 | 0.084 |
| Neg. Cont. 6 | N/A | 0.012 | 0.001 |
| Neg. Cont. 7 | N/A | 0.142 | 0.001 |

**Supplementary Table 4.** SARS-CoV-2 prevalence (percent of cases per day) for each collection day in cohorts 1 and 2 in the study.

| Collection day | Total Samples | Positive Cases | % Prevalence |
| --- | --- | --- | --- |
| 28-May | 41 | 0 | 0.000 |
| 2-Jun | 98 | 0 | 0.000 |
| 3-Jun | 86 | 0 | 0.000 |
| 4-Jun | 108 | 0 | 0.000 |
| 9-Jun | 123 | 0 | 0.000 |
| 10-Jun | 134 | 0 | 0.000 |
| 11-Jun | 141 | 0 | 0.000 |
| 23-Jun | 203 | 1 | 0.493 |
| 24-Jun | 216 | 1 | 0.463 |
| 25-Jun | 314 | 3 | 0.955 |
| 30-Jun | 121 | 2 | 1.653 |
| 1-Jul | 115 | 0 | 0.000 |
| 2-Jul | 102 | 1 | 0.980 |

